# Correcting drug-resistance prevalence in tuberculosis using a predictive model of diagnostic access

**DOI:** 10.1101/2025.10.27.25338839

**Authors:** Jhancy Rocio Aguilar-Jimenez, Fredi Alexander Diaz-Quijano

**Affiliations:** University of São Paulo, School of Public Health, Postgraduate Program in Epidemiology, Laboratory of Causal Inference in Epidemiology (LINCE-USP), Postal address: Av. Dr. Arnaldo 715 – Cerqueira César, São Paulo-SP, 01246-904, Brazil; Universidad de Santander, Facultad de Ciencias Médicas y de la Salud, Bucaramanga, Colombia; University of São Paulo, School of Public Health, Department of Epidemiology, Laboratory of Causal Inference in Epidemiology (LINCE-USP), Postal address: Av. Dr. Arnaldo 715 – Cerqueira César, São Paulo-SP, 01246-904, Brazil

**Keywords:** Drug-resistant, tuberculosis, *Xpert*^®^*MTB/RIF*, *Mycobacterium tuberculosis*, diagnosis, Brazil

## Abstract

**Background:** Drug resistance (DR) poses a major challenge to tuberculosis (TB) elimination. Incomplete access to diagnostic tests can lead to under detection and biased DR-TB prevalence indicators. This study aimed to estimate the prevalence of DR-TB, identified through any available diagnostic method, while correcting for disparities in access to rapid molecular testing (*Xpert*^®^*MTB/RIF*), used as a proxy for overall diagnostic capacity.

**Methods:** We conducted a cross-sectional study of new TB cases in individuals aged over 18 (n=406,331), using 2015-2020 data from the Brazilian Information System on Notifiable Diseases (SINAN). We assumed that access to *Xpert*^®^*MTB/RIF* was an indicator of better diagnostic conditions for detecting drug resistance in general, because patients who underwent this test had a higher prevalence of all forms of resistance (not just to rifampicin). We developed a multilevel mixed-effects logistic regression model, incorporating individual- and municipality-level variables, to predict access to *Xpert*^®^*MTB/RIF* testing in a random sample of 81,027 observations. The model was validated using out-of-sample observations across geographic areas. We then used inverse probability weighting based on predicted access to calculate corrected estimates of DR-TB prevalence.

**Results:** The model showed good performance (AUC=80·93%, 95%CI: 80·71%-81·33%). The prevalence of DR-TB reported by the surveillance system was 1·54% (95%CI: 1·50%-1·57%). Among patients tested with *Xpert*^®^*MTB/RIF*, DR-TB prevalence was 3·86% (95%CI: 3·75%-3·96%), and the corrected weighted prevalence was 5·95% (95%CI: 5·54%-6·38%).

**Conclusions:** The prevalence of DR-TB may be substantially underestimated due to uneven access to testing. Our approach highlights the importance of correcting for diagnostic access to improve surveillance indicators.

## Background

Global progress toward meeting the goals of the End TB Strategy has slowed down in recent years.^1^ The increase in drug-resistant tuberculosis (DR-TB) remains a public health crisis^2^ and it is estimated that, worldwide, almost half a million people will develop multidrug-resistant or rifampicin-resistant TB (MDR/RR-TB) each year.^3^ Underdiagnosis and under-reporting remain significant challenges for the timely treatment and TB transmission control.^4–7^ It was estimated that only 44% of global drug-resistant TB cases were diagnosed and reported in 2019.^8^

The World Health Organization (WHO) recommends equitable access to rapid molecular testing for TB diagnosis.^9^ In 2021, only 38% of new TB cases worldwide had access to a WHO-recommended rapid diagnostic test (WRD) as an initial test and merely a quarter of diagnostic centers worldwide were equipped to perform these tests.^10^ Brazil remains one of the countries with the highest number of TB cases^4^ and accounts for 33·4% of the disease burden in the Americas.^11^ While *Xpert*^®^*MTB/RIF* testing is established for diagnosing pulmonary and extrapulmonary TB,^12^ only 30·7% of diagnoses of new cases reported between 2015 and 2020 in the Brazilian epidemiological surveillance system were made using this method.^13^ Non-universal access to this and other diagnostic methods may bias the estimated measures. For instance, in Brazil, the epidemiological surveillance system reports only 1·5% of drug-resistant TB (DR-TB) ^13^, which is considerably lower than findings from other studies employing systematic diagnostic approaches, where prevalence ranges from 6.1% for MDR-TB to 13·6% for resistance to at least one antibiotic among previously untreated patients from certain populations in Brazil. ^14^ Consequently, the limited availability of diagnostic tools could explain this underreporting, given that only a minority of patients with suspected MDR-TB have access to antimicrobial susceptibility testing.^15^

Drug resistance can be identified through various diagnostic methods, such as culture-based drug susceptibility testing and molecular assays. However, information on access to many of these tools is either not systematically recorded in surveillance data or depends on prior steps (such as obtaining a positive culture result), which may not be uniformly available. In contrast, access to *Xpert®MTB/RIF* testing is explicitly documented in the Brazilian TB surveillance system, and the test can be performed directly on sputum samples, independently of prior test results, making it a more reliable and direct indicator. While *Xpert*^®^*MTB/RIF* testing only evaluates the rifampicin resistance profile, the implementation of this test has been associated with an increase in the sensitivity of the surveillance system to identify drug resistance in general.^13,16^

Since the determinants of access to diagnostic testing may also be associated with DR-TB, we aimed to identify and use them to correct for selection bias in prevalence estimates. Accordingly, using national epidemiological data, we developed a predictive model of access to *Xpert*^®^*MTB/RIF* testing, considered a proxy for overall diagnostic capacity. We then applied inverse probability weighting based on this model, to obtain corrected DR-TB prevalence estimates from surveillance data.

## Materials and methods

### Study design and source of data

This cross-sectional study analyzed secondary data from the Brazilian Information System on Notifiable Diseases (SINAN), to estimate the prevalence of DR-TB while correcting for unequal access to diagnostic tests. This information system is utilized for the mandatory notification of all new TB cases diagnosed in the country and was estimated to have a reported case detection rate of 87%.^17^

Data on new TB cases in individuals over 18 years old, reported between 01/01/2015 and 12/31/2020, were extracted anonymously from SINAN on 01/14/2022. Thus, all included patients had been diagnosed with TB at least one year prior to the data extraction. This ensured that the classification of drug resistance was updated in the surveillance system, as some cases require confirmation of the diagnosis of DR-TB. Data from 2014 or earlier were not included, because this was the year of implementation of the *Xpert*^®^*MTB/RIF* system in Brazil. Pediatric TB cases were omitted because of the test’s limited use in this population, due to sputum collection challenges and low bacilli presence.^18^ Contextual variables for 4,998 municipalities, which reported new TB cases during the study period, were obtained from the Brazilian Ministry of Health and the SINAN routine TB surveillance database in 2013.

Given that access to *Xpert*^®^*MTB/RIF* is systematically recorded in the SINAN database, we used it as a proxy for broader diagnostic capacity to detect drug resistance in general. We consider this assumption valid, as we observed that among those with access to this diagnostic method, there was a higher frequency of other forms of resistance (not only to rifampicin), suggesting a generally higher sensitivity for detecting TB-DR cases. Specifically, resistance prevalences to isoniazid, rifampicin, both drugs, and other first-line agents increased from 0·35%, 0·74%, 0·18%, and 0·27% in the general TB population to 0·71%, 2·27%, 0·37%, and 0·50%, respectively, among individuals with access to *Xpert*^®^*MTB/RIF*. Based on these findings, we assumed that *Xpert*^®^*MTB/RIF* testing was associated with increased access to all drug-resistance diagnostic methods. Therefore, we developed a predictive model for access to this test to correct potential selection bias in DR-TB prevalence estimates.

### Study variables

Access to *Xpert*^®^*MTB/RIF* was analyzed as a dichotomic variable. Cases registered in the database classified as “Rifampicin-sensitive detectable TB”, “Rifampicin-resistant detectable TB”, “undetectable”, and “inconclusive” were considered cases with access to *Xpert*^®^*MTB/RIF* testing. Cases in which this test was not performed, ignored, or not completed were grouped in the no-access category.

Individual variables interpreted as characteristics preceding the diagnosis were considered potential covariates of the predictive model. Sociodemographic variables included sex, race, age, education, beneficiary of governmental income transfer programs, population deprived of liberty, homeless, health professionals and immigrants. Other clinical predictors included AIDS, HIV, drug use, smoking, alcoholism, diabetes, mental illness, and form of the disease (pulmonary, extrapulmonary or mixed).

We considered two municipal contextual variables: (1) the availability of the *Xpert*^®^*MTB/RIF* system (a dichotomous variable), and (2) the proportion of tuberculosis (TB) cases with drug resistance in 2013, as detected by any of the available sensitivity tests.

### Statistical analysis

Database was exported to Stata® 18·0 software (Stata Corp LP, College Station, TX, USA) for analysis. Demographic and clinical characteristics were described using proportions for qualitative variables and medians and interquartile range for quantitative variables. The most functional form of quantitative variables, such as age, was sought, evaluating their linear relationship to the dependent variable.

We used a mixed-effects multilevel logistic regression for the predictive model of access to *Xpert*^®^*MTB/RIF* testing. A non-automatic stepwise logistic regression procedure was used for covariate selection. In a preliminary analysis, variables not associated with outcome (p > 0·2) were excluded from the analysis. To evaluate multiplicative interactions, we created and tested 199 interaction terms with each of the independent predictors. Variables with p-values lower than 0·05 were retained in the final multiple model. To choose the final model, criteria such as the Akaike information criterion (AIC) and the Bayesian information criterion (BIC) ^19^ were assessed, as well as the pseudo R2.

The model was obtained using a random sample of 81,027 observations, of which 31·0% had access to *Xpert*^®^*MTB/RIF* (the dependent variable). Validation was performed using observations not included in the development of the predictive model, covering seven geographic areas: North (37,598 cases), Northeast (83,921), Southeast (19,948), South (39,387), Midwest (15,637), Rio de Janeiro (49,801), and São Paulo (78,856). Rio de Janeiro and São Paulo states were analyzed separately due to their high case concentrations.

As a measure of discrimination, we calculated the area under the receiver operating characteristic (AUC-ROC) curve. We considered values between 0·7 and 0·8 as acceptable, between 0·8 and 0·9 as good and above 0·9 as excellent.^20^ To evaluate the calibration, we compared the number of observed and predicted values by deciles and calculated the Brier score.

### DR TB prevalence estimates

A case of DR-TB was defined as a positive result from any drug resistance diagnostic method and was therefore not limited to rifampicin resistance alone. Drug resistance prevalence in TB patients was calculated using the following three approaches:

1. Prevalence of DR-TB recorded in the surveillance system (unweighted): corresponded to the estimated prevalence of resistance detected by any method in all patients, according to the official records of the surveillance system.
2. Unweighted prevalence of DR-TB restricted to patients with access to the *Xpert*^®^*MTB/RIF* testing: this refers to drug resistance detected by any method, but only in the group of patients with access to the *Xpert*^®^*MTB/RIF* testing.
3. Weighted prevalence of DR-TB, which refers to the prevalence of drug resistance identified by any method among TB patients who had access to *Xpert*^®^*MTB/RIF* testing, with these observations weighted by the inverse of the probability of access to the test, according to the predictive model. In this way, the resulting estimate is expected to reflect the overall DR-TB prevalence under improved sensitivity conditions, considering that it is based on cases with access to *Xpert*^®^*MTB/RIF* testing and, at the same time, represents the entire population due to the weighting procedure. This third weighted estimate was used as a reference to calculate the underreporting of the Surveillance System.

Specifically:

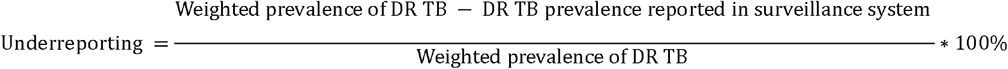

### Ethics Statement

This study used publicly available data and did not include personal information of TB patients. According to Resolution no. 510 of the Brazilian National Health Council, this type of research does not require evaluation by an institutional review board.

## Results

Between 2015 and 2020, 406,331 new TB cases in patients over 18 were reported in Brazil. Of these, 69·6% were men, 65·2% lived in urban areas, 56·3% were black/brown, and 59·8% were aged 25-54. The proportion of access to *Xpert*^®^*MTB/RIF* testing varied geographically, from 37·7% (95% CI: 37·36%-37·96%) in São Paulo to 25·1% (95%CI 24·9% - 25·4%) in the Northeast region (Table 1).

**Table 1.** Frequency of access to *Xpert*^®^*MTB/RIF* testing according to the individual characteristics of new cases of tuberculosis in the population aged 18 years and over in Brazil, 2015-2020 (n=406,331)

|  | TB cases |  | With access *Xpert*<sup>®</sup> MTB/RIF |  |
| --- | --- | --- | --- | --- |
|  | n total | n | % | 95% CI |
| **Brazil (consolidated)** | 406331 | 126466 | 31.12 | (30.98 – 31.27) |
| **Geographical areas** |  |  |  |  |
| North | 47022 | 15706 | 33.4 | (32.98 - 33.83) |
| Northeast | 104848 | 26329 | 25.11 | (24.85 - 25.37) |
| Southeast | 24951 | 7304 | 29.27 | (28.71 – 29.84) |
| South | 49220 | 13837 | 28.11 | (27.72 - 28.51) |
| Central-West | 19454 | 5178 | 26.62 | (26 - 27.24) |
| Rio de Janeiro State | 62250 | 20991 | 33.72 | (33.35 - 34.09) |
| São Paulo State | 98381 | 37052 | 37.66 | (37.36 - 37.96) |
| **Sex** |  | -- | -- | -- |
| Female | 123698 | 35113 | 28.39 | (28.13 - 28.64) |
| Male | 282604 | 91346 | 32.32 | (32.15 - 32.49) |
| **Aged** | 406331 | -- | -- | -- |
| 18 - 24 | 70113 | 23433 | 33.42 | (33.07 - 33.77) |
| 25 - 34 | 97932 | 32798 | 33.49 | (33.2 - 33.79) |
| 35 - 54 | 145212 | 44829 | 30.87 | (30.63 - 31.11) |
| ≥55 | 93074 | 25406 | 27.3 | (27.01 - 27.58) |
| **Form** |  | -- | -- | -- |
| Pulmonary | 341407 | 115113 | 33.72 | (33.56 - 33.88) |
| Extrapulmonary | 52721 | 7315 | 13.87 | (13.58 - 14.17) |
| Pulmonary+Extrapulmonary | 12024 | 4038 | 33.58 | (32.74 - 34.44) |
| **Race/ethnicity** |  | -- | -- | -- |
| White | 112005 | 31534 | 28.15 | (27.89 - 28.42) |
| Black | 46490 | 15234 | 32.77 | (32.34 - 33.20) |
| Asian descent | 2432 | 778 | 31.99 | (30.14 - 33.89) |
| Brown ("Parda") | 182183 | 56544 | 31.04 | (30.82 - 31.25) |
| Indigenous | 3531 | 840 | 23.81 | (22.43 - 25.24) |
| Ignored | 59690 | 21536 | 36.08 | (35.69 - 36.47) |
| **Area** |  | -- | -- | -- |
| Urban | 264871 | 79106 | 29.87 | (29.69 - 30.04) |
| Rural | 31000 | 6710 | 21.65 | (21.19 - 22.11) |
| Peri-urban | 2852 | 981 | 34.40 | (32.65 - 36.17) |
| Ignored | 107608 | 39669 | 36.86 | (36.58 - 37.15) |
| **Special populations** | -- | -- | -- | -- |
| Prisoners | 44191 | 18331 | 41.48 | (41.02 – 41.94) |
| Migrant population | 2593 | 1135 | 43.77 | (41.85 - 45.70) |
| Health-care workers | 6006 | 1820 | 30.30 | (29.14 - 31.48) |
| Homeless people | 10651 | 5067 | 47.57 | (46.62 - 48.53) |
| **Specific condition** | -- | -- | -- | -- |
| Illicit drug use | 51939 | 21913 | 42.19 | (41.76 - 42.62) |
| Tobacco smoking | 90693 | 34086 | 37.58 | (37.27 - 37.90) |
| Alcohol abuse | 69970 | 24471 | 34.97 | (34.62 - 35.33) |
| AIDS | 35204 | 11926 | 33.88 | (33.38 - 34.37) |
| Diabetes | 34275 | 10741 | 31.34 | (30.85 - 31.83) |
| Mental illness | 9265 | 2670 | 28.82 | (27.9 - 29.75) |
| **Outcome classification** | -- | -- | -- | -- |
| Cure | 278473 | 86637 | 31.11 | (30.94 - 31.28) |
| Abandonment | 44448 | 15645 | 35.2 | (34.76 - 35.64) |
| Death | 29609 | 7269 | 24.55 | (24.06 - 25.04) |

Access to this rapid molecular test by area of residence was higher among peri-urban residents (34·4%) compared to urban (29·9%) and rural areas (21·7%). Among special populations, homeless individuals (47·6%) had the greatest access, followed by prisoners (41·5%), migrants (43·8%), and healthcare workers (30·3%). Illicit drug users (42·2%) had higher access compared to people with AIDS (33·9%) and those with alcohol abuse (35.0%) (Table 1).

A multiple model integrated four sociodemographic variables (age, sex, race/ethnicity, and educational level); three special conditions (smoking, drug use, and AIDS); three special populations (prisoners, homeless individuals, and migrants); 15 interaction terms; and the contextual predictors, *Xpert*^®^*MTB/RIF* system availability and municipal drug-resistance proportion from 2013 (Table 2). The model’s AUC was 80·9% (95% CI: 80·7%-81·3%) in the specification data set. In the validation data set, the lowest AUC corresponded to the state of Rio de Janeiro (74·3%; 95% CI: 73·8%-74·7%). In the other geographic areas, it ranged from 78·2% to 84·8% (Fig 1).

**Table 2.** Multilevel mixed-effects predictive model for access to *Xpert*^®^*MTB/RIF* testing for TB diagnosis in the population aged 18 years and over in Brazil, 2015-2020 (n=406,331)

| Variables | OR adjusted (95% CI) | p-valued |
| --- | --- | --- |
| **Individual level** |  |  |
| **Sex** | -- | -- |
| Female | Ref | -- |
| Male | 1.18 (1.11 - 1.25) | <0.001 |
| **Race/ethnicity** | -- | -- |
| Other race/ethnicity | Ref | -- |
| Black and brown ("Parda") | 1.22 (1.14 - 1.31) | <0.001 |
| **Educational level** | -- | -- |
| Up to 4th grade of primary schools | Ref | -- |
| 5th grade incomplete - High school incomplete | 1.01 (0.96 - 1.07) | 0.636 |
| High school completed | 0.86 (0.80 - 0.93) | <0.001 |
| Higher education incomplete or complete | 0.67 (0.62 - 0.73) | <0.001 |
| Ignored | 0.72 (0.68 - 0.77) | <0.001 |
| **Tobacco smoking** | 1.27 (1.2 - 1.34) | <0.001 |
| **Illicit drug use** | -- | -- |
| No | Ref | -- |
| Yes | 1.38 (1.27 - 1.5) | <0.001 |
| Ignored | 0.72 (0.63 - 0.84) | <0.001 |
| **AIDS** | -- | -- |
| No | Ref | -- |
| Yes | 0.88 (0.83 - 0.94) | <0.001 |
| Ignored | 0.81 (0.74 - 0.89) | <0.001 |
| **Prisoners** | 1.31 (0.95 - 1.82) | 0.103 |
| **Homeless population** | -- | -- |
| No | Ref | -- |
| Yes | 1.47 (1.24 - 1.74) | <0.001 |
| Ignored | 0.62 (0.52 - 0.73) | <0.001 |
| **Migrant population** | 1.51 (1.25 - 1.83) | <0.001 |
| **Ln (diagnosis year)** | 1.89 (1.83 - 1.95) | <0.001 |
| **Ln (age) (in years)** | 8.26 (3.46 - 19.71) | <0.001 |
| **Ln (age\*age)** | 0.74 (0.65 - 0.83) | <0.001 |
| **Interactions terms** | -- | -- |
| Sex*race | 0.91 (0.84 - 0.98) | 0.012 |
| Sex*prisoners | 1.43 (1.05 - 1.95) | 0.024 |
| Race/ethnicity*prisoners | 0.78 (0.70 - 0.88) | <0.001 |
| Race/ethnicity*migrant | 1.33 (1.12 - 1.56) | 0.001 |
| Tobacco smoking*drug | 0.83 (0.74 - 0.92) | <0.001 |
| Tobacco smoking*education | 1.14 (1.03 - 1.25) | 0.008 |
| Tobacco smoking*prisoners | 0.37 (0.3 - 0.46) | <0.001 |
| Drug*prisoners | 0.83 (0.72 - 0.95) | 0.008 |
| Drug*homeless | 0.75 (0.61 - 0.93) | 0.007 |
| Drug*AIDS | 1.31 (1.11 - 1.54) | 0.002 |
| AIDS*prisoners | 0.47 (0.39 - 0.57) | <0.001 |
| AIDS*education | 1.24 (1.08 - 1.42) | 0.002 |
| Prisoners*education | 1.19 (1.05 - 1.36) | 0.007 |
| Homeless*education | 1.32 (1.07 - 1.62) | 0.009 |
| Prisoners*Ln(diagnosis year) | 1.27 (1.15 - 1.39) | <0.001 |
| **Municipality level** |  |  |
| ***Xpert*<sup>®</sup> *MTB/RIF* system availability** | 5.70 (4.41 - 7.38) | <0.001 |
| **Previous resistance proportion (each percentage point)** | 1.07 (1.03 - 1.12) | 0.001 |

**Figure 1.**
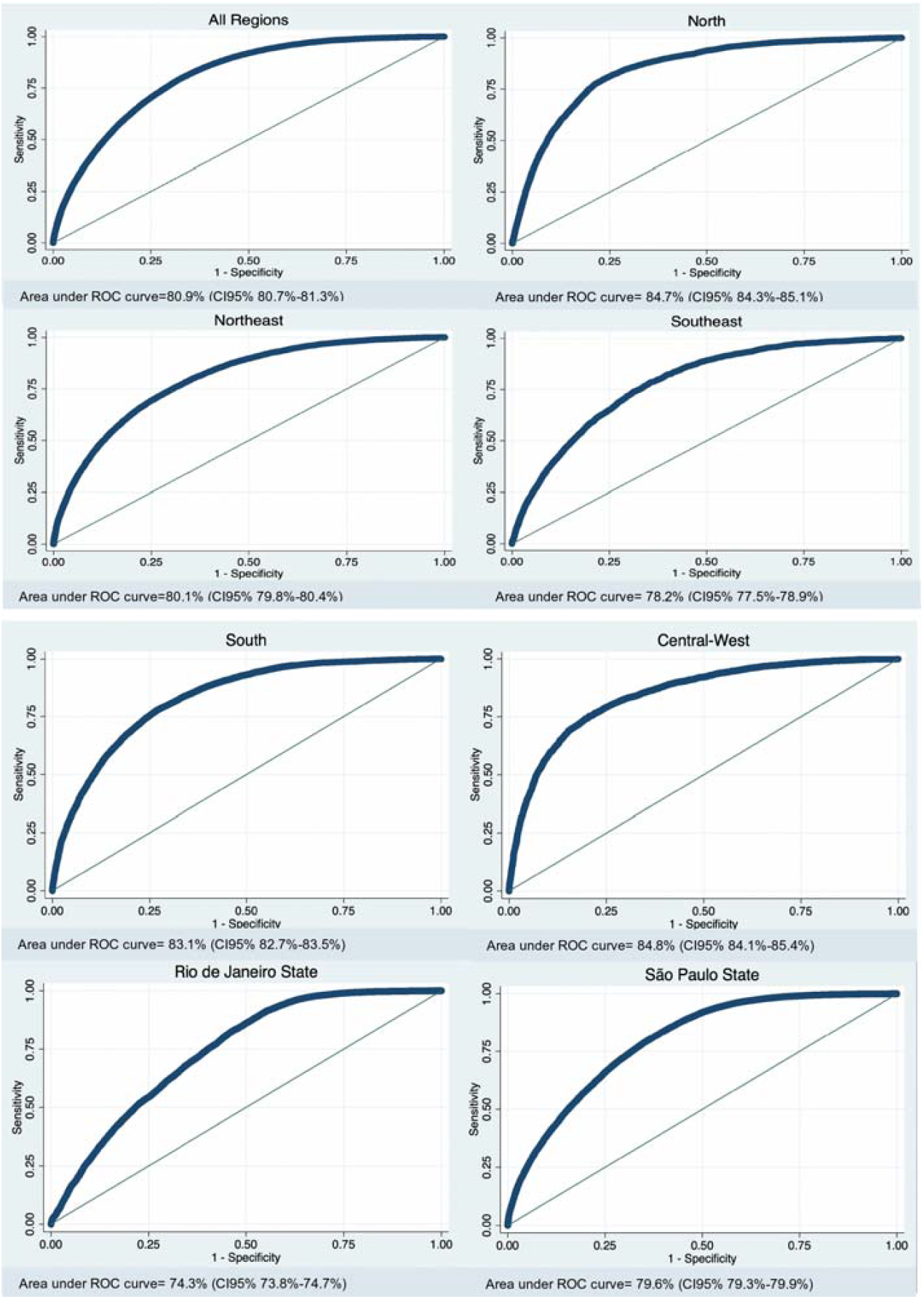
Discrimination of the multilevel predictive model by estimating the areas under the ROC curve for the different Brazilian geographic areas.

Comparing the observed cases with those predicted by the model, we considered that there was adequate model calibration for most geographic areas. The Brier score values were less than 0·19. The only geographic area where there was an appreciable gap between observed and predicted cases was the Central West region; however, the Brier score for this region was 0·13, which is lower than in other regions (Fig 2).

**Figure 2.**
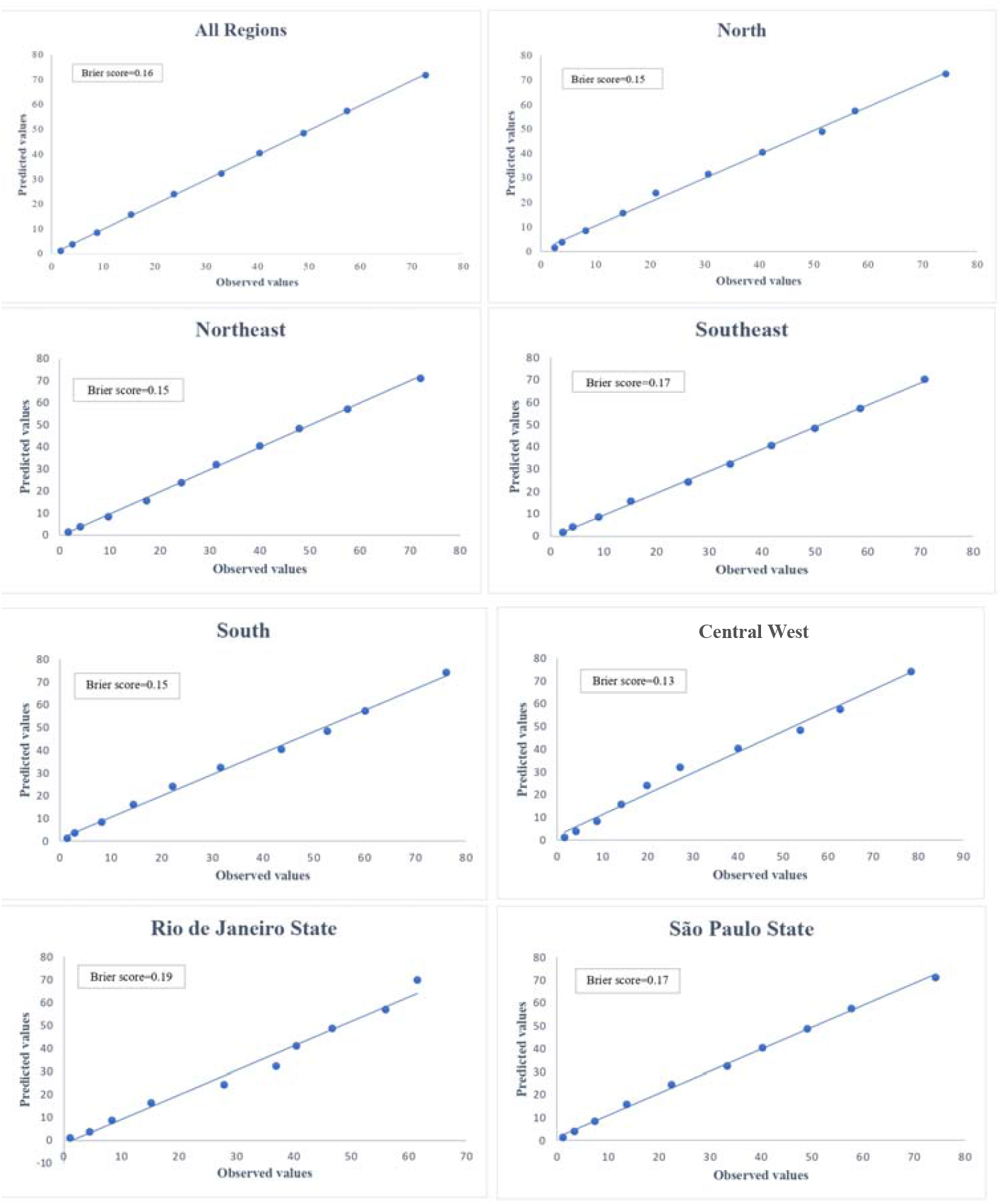
Calibration plots of the multilevel prediction model for each geographic area of Brazil.

The prevalence of DR-TB reported by the surveillance system was 1·54% (95% CI: 1·50%-1·57%). Among the group with access to *Xpert*^®^*MTB/RIF* testing, the prevalence of DR-TB was 3·86% (95% CI: 3·75%-3·96%). When prevalence was weighted, it increased to 5·95% (95% CI: 5·54%-6·38%). This trend, where the weighted estimate is higher than other prevalence estimates, was consistent across geographic areas. This aligns with the distribution of DR-TB prevalence according to predicted values of access to *Xpert*^®^*MTB/RIF* testing, revealing a pattern of higher drug resistance prevalence in deciles with lower probabilities of *Xpert*^®^*MTB/RIF* access (Fig 3).

**Figure 3.**
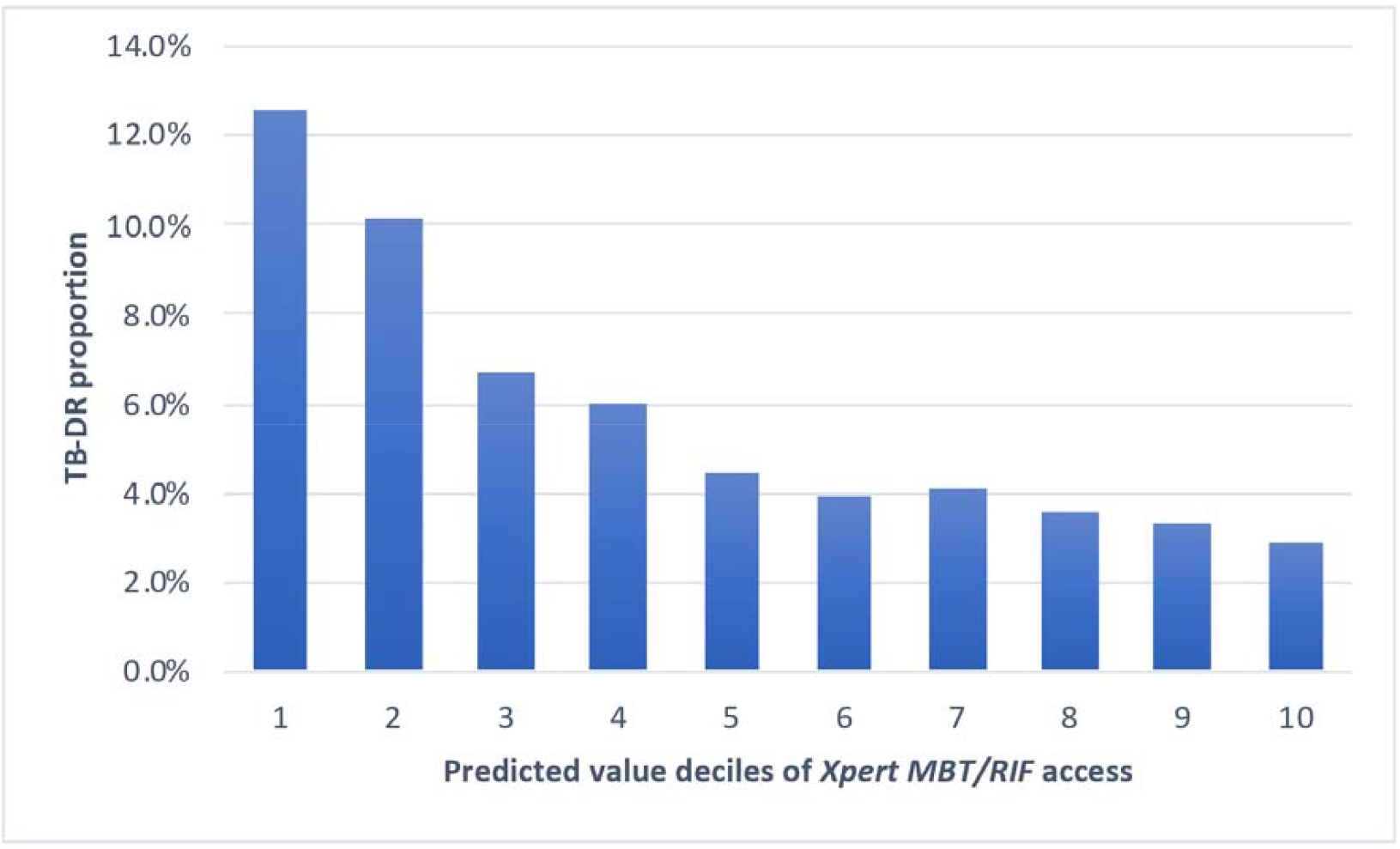
Prevalence of DR-TB in patients who had access to *Xpert*^®^*MTB/RIF* testing by decile of predicted value.

When comparing the geographic areas, the highest weighted prevalence of DR-TB was observed in the South region (8·77%; 95% CI: 7·43%-10·33%), followed by the state of Rio de Janeiro (7·18%; 95% CI: 5·80-8·87) and the Northeast region (7·12%; 95% CI: 6·46-7·84). The lowest weighted prevalence of DR-TB was recorded in the state of São Paulo (3·16%; 95% CI: 2·52-3·96). Furthermore, the weighted prevalence of DR-TB of 8·43% (95% CI: 7·04% - 10·07%) in the rural area corresponded to almost four times that recorded in the peri-urban area (2·27%; 95% CI: 1·12-4·53; see Table 3).

**Table 3.**
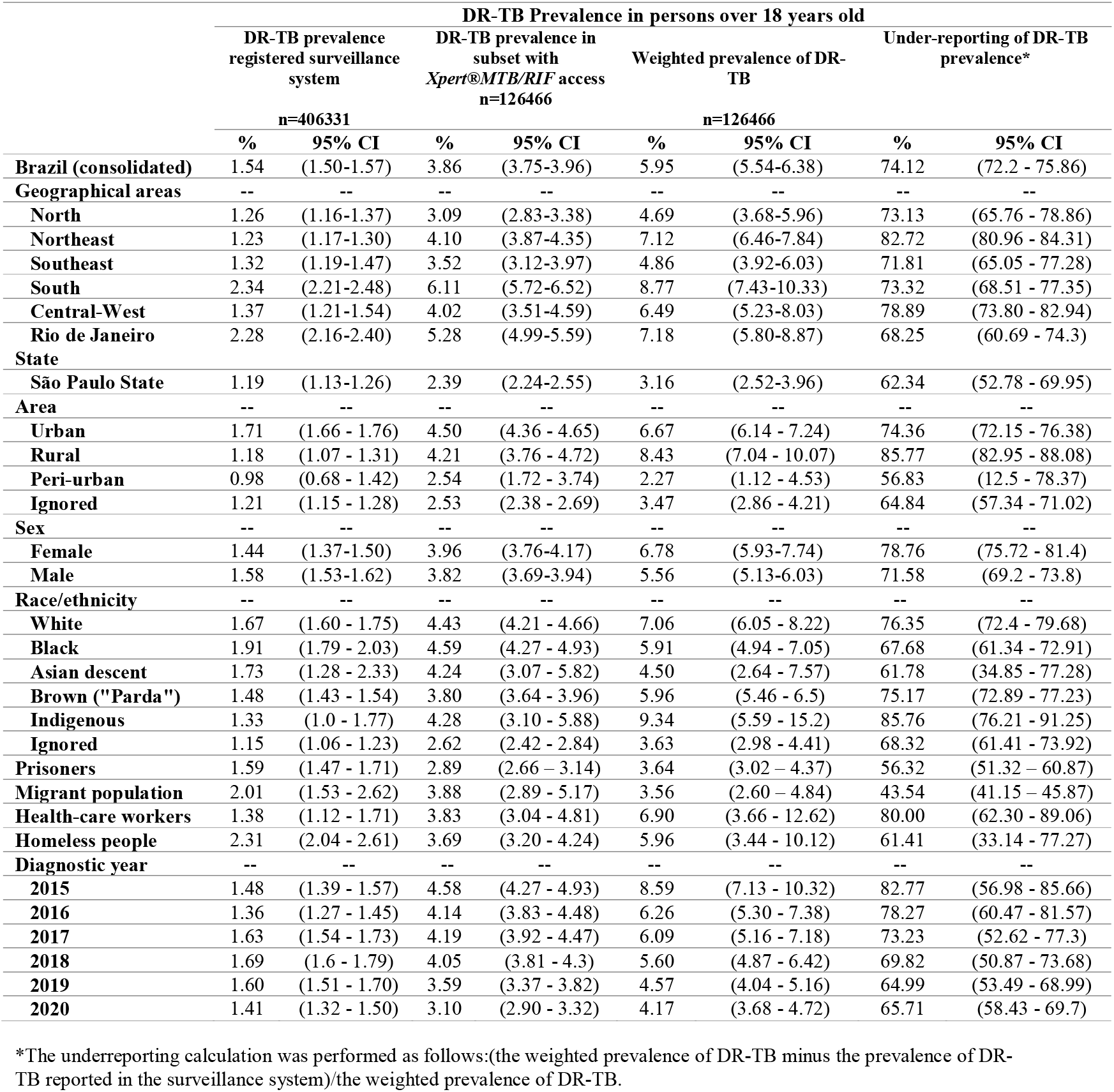
Comparison of the prevalence of DR-TB between the surveillance system, *Xpert*^®^*MTB/RIF* and weighted estimates for the general population over 18 years of age in Brazil.

The weighted prevalence of DR-TB was higher in females (6·78%; 95%CI 5·93-7·74) than males (5·56%; 95%CI 5·13-6·03). By ethnicity, indigenous populations had the highest prevalence (9·34%; 95%CI 5·59% - 15·2%), while those of Asian descent had the lowest (4·50; 95%CI 2·64% - 7·57%). Among special populations, healthcare workers had the highest weighted prevalence of DR-TB (6·90%; 95%CI 3·66% - 12·62%), followed by the homeless (5·96%; 95%CI 3·44% - 10·12%), prisoners (3·64%; 95%CI 3·02% - 4·37%) and immigrants (3·56%; 95%CI 2·60% - 4·84%; see Table 3).

We estimated that the surveillance system had an underreporting of DR-TB prevalence of 74·12% (95%CI 72·2% - 75·86%). The geographic area with the highest under-reporting was the Northeast region (82·72%; 95%CI 80·96% - 84·31%), followed by the Central-West region (78·89%; 95%CI 73·80% - 82·94%) and the lowest value was recorded in the state of São Paulo (62·34%; 95%CI 52·78% - 69·95%). According to area, the rural area presented the highest underreporting of DR-TB with values of 85·77% (95%CI 82·95% - 88·08%) and the peri-urban area the lowest (56·83%; 95%CI 12·5% - 78·37%; see Table 3).

Both women (78·76%; 95%CI 75·72% - 81·4%) and men (71·58%; 95%CI 69·2% - 73·8%) showed DR-TB prevalence underreporting above 70%. Among ethnic groups, underreporting was highest in indigenous populations (85·76%; 95%CI 76·21% - 91·25%) and whites (76·35%; 95%CI 72·4% - 79·68%). When comparing special populations, healthcare workers had the highest underreporting (80%; 95%CI: 62·30% - 89·06%), while immigrants had the lowest (43·54%; 95%CI: 41·15% - 45·87%). Underreporting progressively decreased from 82·77% (95% CI: 56·98% - 85·66%) in 2015 to 64·99% (53·49%-68·99%) in 2019, with a slight increase in 2020 (65·71%; 95% CI: 58·43% - 69·7%; see Table 3).

## Discussion

Restricting the analysis to the group with access to *Xpert*^®^*MTB/RIF* testing is expected to increase the sensitivity for identifying drug resistance in general ^13^, but this group does not represent the entire population and must be appropriately weighted. In this regard, we developed a predictive model of *Xpert*^®^*MTB/RIF* testing access, incorporating contextual and individual variables. The inverse probability weights derived from this model were used to control for selection biases related to access to diagnostic technology, which may also be associated with the likelihood of drug resistance.

Although *Xpert*^®^*MTB/RIF* detects only rifampicin resistance, we assumed that access to this test reflects better diagnostic conditions overall, such as greater availability of laboratory infrastructure, trained personnel, and diagnostic follow-up, which would increase the chances of detecting drug resistance by any available method. Accordingly, we consider that weighted estimates provide a more accurate representation of DR-TB prevalence in the population.

We consider that the validation of the model was satisfactory across the different geographic areas. Even in the state of Rio de Janeiro, where the model exhibited the lowest discrimination, the value of the AUC-ROC exceeded 70%. Although variations in the predictors may have slightly affected the accuracy, it remained within levels that we consider acceptable.

All population groups had weighted DR-TB estimates higher than the official estimates, recorded by the epidemiological surveillance system. These weighted estimates were even higher than those obtained by observing only the group with access to *Xpert*^®^*MTB/RIF* testing. Our estimate of DR-TB underreporting in the country (74%) was higher than that previously suggested by the WHO (67%).^21^ Our correction of the DR-TB prevalence results in values more consistent with those reported in studies employing systematic and rigorous drug resistance assessment.^14,23^ The advantage of our implemented method is that it can be applied on a large scale using epidemiological surveillance data, allowing for continuous monitoring of trends and their determinants.

Comparing the different geographic areas of the country, the Northeast region showed the highest underreporting, which aligns with previous studies reporting limited access to diagnostic tests in this area.^22^ Populations in remote areas, such as indigenous and rural communities, showed higher underreporting of DR-TB cases. In Brazil, socioeconomic conditions, including poor housing, malnutrition, inadequate work, and limited healthcare access, worsen in remote regions.^24^

Additionally, primary healthcare in indigenous villages has been found inefficient in detecting respiratory symptoms.^25^ In this way, our findings align with the recent WHO recommendation to improve access to diagnostic test in peripheral, high-risk, and marginalized areas. Access, however, may still be limited by local policies, diagnostic algorithms or restrictions on the use of WRD to selected populations.^26^

Methodologically, the change in prevalence estimates after applying the weighting correction is notable. While we expected an increase in DR-TB prevalence among patients with access to *Xpert*^®^*MTB/RIF*, the weighted estimate exceeded this group’s prevalence. This result becomes intuitive when considering the negative association observed between test access and resistance. As illustrated in Fig 3, DR-TB prevalence decreases in categories with higher probabilities of test access, explaining why the weighted values surpass the estimates based solely on those with access to *Xpert*^®^*MTB/RIF* testing. We believe this highlights the importance of weighting to correct for biases arising from complex relationships between the determinants of access and resistance.

On the one hand, demographic, clinical and geographic differences between people with and without access pose two major problems: 1) inequity and 2) biases in estimating the prevalence of DR-TB. Efforts must be intensified to achieve universal diagnosis of TB and drug resistance, particularly in high-burden countries where drug susceptibility testing lacks universal coverage.^4,27^ The second phase of the National Plan to End TB emphasizes prevention and integrated, patient-centered care, including active case finding and promoting early diagnosis of both sensitive and resistant TB, with timely treatment initiation.^11^

Our study had certain limitations that may affect the generalizability of the results. Although we used access to *Xpert*^®^*MTB/RIF* testing as a proxy for adequate diagnostic conditions, this test only detects rifampicin resistance and requires complementary methods to assess resistance to other drugs. Nevertheless, as previously shown,^13^ its availability is a reasonable indicator of improved sensitivity for detecting DR-TB, especially since it is often used in combination with other diagnostic tools.

Regarding the predictive model for access to *Xpert*^®^*MTB/RIF* testing, it achieved good discrimination and adequate calibration across most geographic areas. The Central-West region was an exception, showing discrepancies between predicted and observed values. However, considering this region’s low TB case numbers and moderate DR-TB prevalence, we anticipate minimal impact on overall estimates.

Estimating the underreporting of DR-TB prevalence can be challenging. Some studies have opted to use active case finding or drug dispensing, which could be considered more accurate methods for quantifying the true frequency of DR-TB.^28^ However, such approaches are not feasible on a large scale. Despite its limitations, the approach proposed in this study could facilitate the monitoring of resistance underreporting at a broader level.

In conclusion, we obtained a predictive model for access to the *Xpert*^®^*MTB/RIF* testing, used as a proxy for access to drug resistance diagnostic tools. The model included individual and contextual variables and exhibited good performance across different geographic areas of Brazil. This model was used to weight the observations, allowing DR-TB prevalence estimates to be corrected for biases attributable to differential access to diagnostic tools. As a result, we observed a much higher underreporting of DR-TB than previously estimated by the WHO, suggesting that at least three-quarters of DR-TB cases were not identified in Brazil.

## Data Availability

The data supporting the findings of this study are not publicly available but can be accessed upon reasonable request to the Brazilian Ministry of Health (https://www.gov.br)

## Data and Code Availability Statement

Researchers can access the datasets by navigating to the health statistics section and selecting the appropriate data files. All data used were anonymized and aggregated, in compliance with relevant data protection regulations.

## Author Contributions

All authors contributed to the study conception and design. Material preparation, data collection and analysis were performed by Jhancy R. Aguilar-Jiménez and Fredi A. Diaz-Quijano. The first draft of the manuscript was written by Jhancy R. Aguilar-Jiménez and all authors commented on previous versions of the manuscript. All authors read and approved the final manuscript.

## List of abbreviations

DR: Drug resistance
TB: tuberculosis
SINAN: Brazilian Information System on Notifiable Diseases
DR-TB: drug-resistant tuberculosis
95% CI: 95% Confidence Interval
MDR-TB: multidrug-resistant
RR-TB: rifampicin-resistant TB
WHO: World Health Organization
WRD: WHO-recommended rapid diagnostic test
AUC-ROC: area under the receiver operating characteristic curve.

